# Mental health, substance use and viral load suppression in adolescents receiving ART at a large paediatric HIV clinic in South Africa

**DOI:** 10.1101/2020.07.06.20147298

**Authors:** Andreas D. Haas, Karl-Günter Technau, Shenaaz Pahad, Kate Braithwaite, Mampho Madzivhandila, Gillian Sorour, Shobna Sawry, Nicola Maxwell, Per von Groote, Mpho Tlali, Mary-Ann Davies, Matthias Egger, for the IeDEA Southern Africa Collaboration

**Affiliations:** Institute of Social & Preventive Medicine, University of Bern, Bern, Switzerland; Empilweni Services and Research Unit, Department of Paediatrics & Child Health, Rahima Moosa Mother and Child Hospital, Faculty of Health Sciences, University of the Witwatersrand, Johannesburg, South Africa; Wits Reproductive Health and HIV Institute, Faculty of Health Sciences, University of the Witwatersrand, Johannesburg, South Africa; Centre for Infectious Disease Epidemiology and Research, University of Cape Town, Cape Town, South Africa

**Keywords:** Adolescents, HIV, mental health screening, depression, viral load suppression, sub-Saharan Africa

## Abstract

**Introduction:** There are few data on the prevalence of mental health problems among adolescents living with HIV in low- and middle-income countries and the evidence on associations between mental health problems and viral load suppression is inconsistent. We assessed the prevalence of mental health problems among adolescents receiving antiretroviral therapy (ART) at a large paediatric HIV clinic in South Africa and examined associations between mental health problems and viral load suppression.

**Methods:** We implemented routine mental health screening at Rahima Moosa Mother and Child Hospital in Johannesburg. Adolescents aged 10-19 years were offered screening for depression (Patient Health Questionnaire-9 [PHQ-9]), suicide (Adolescent Innovations Project [AIP]-handbook), anxiety (General Anxiety Disorder-7 [GAD-7]), trauma (Primary Care PTSD Screen for DSM-5 [PC-PTSD-5]), and substance use (CAGE Adapted to Include Drugs [CAGE-AID]) at each routine HIV care visits. We assessed screening outcomes between February 1, 2018, and January 1, 2020 and calculated odds ratios for associations between positive screening outcomes and unsuppressed viral load (>400 HIV-RNA copies/ml).

**Results:** Out of 1,203 adolescents who attended the clinic, 1,088 (90.4%) were screened at a median age of 13 years (IQR 10-15). In total, 97 (8.9%) screened positive: 48 (4.4%) for depression (PHQ-9 ≥ 10), 29 (2.8%) for suicidal concern, 24 (2.2%) for anxiety (GAD-7 ≥ 10), 38 (3.2%) for trauma (PC-PTSD-5 ≥ 3), and 18 (1.7%) for substance use (CAGE-AID ≥ 2). Positive screening for depression (aOR 2.39, 95% CI 1.02-5.62), trauma (aOR 3.18, 95% CI 1.11-9.07), substance use (aOR 7.13, 95% CI 1.60-31.86), or any mental health condition (aOR 2.17, 95% CI 1.17-4.02) were strongly associated with unsuppressed viral load.

**Conclusions:** HIV-positive adolescents who are affected by mental health or substance use problems are a highly vulnerable population, who merit specific clinical attention. Strategies for screening and management of mental health and substance use problems in adolescents on ART in low- and middle-income countries need to be developed and evaluated.

## Introduction

In 2018, there were an estimated 1.6 million adolescents (aged 10-19 years) living with HIV (ALHIV) globally [1]. The majority of ALHIV live in sub-Saharan Africa, and about 20% in South Africa [1]. ALHIV are a heterogeneous group comprising of those who acquired HIV perinatally and survived into adolescence and those who acquired HIV horizontally [2–4]. In South Africa, the majority of ALHIV enrolled in HIV care acquired HIV perinatally [5].

For a variety of reasons, ALHIV are at risk of developing mental health complications. HIV has a direct effect on the brain, which can lead to neurocognitive decline, behavioral disturbance, mood disorders and psychosis [6]. Furthermore, certain antiretroviral drugs used to treat HIV can have neuropsychiatric side effects [7]. In addition to these biological mechanisms that link HIV and mental illness, several social and structural factors contribute to the poor mental health of ALHIV. Many ALHIV have lost their biological parents and are raised in extended families, or orphanages [8,9]. Caregivers of orphans might be overburdened by the responsibility to care for, and economically support an additional child or children [9]. Orphans often lack social and material support, feel unsafe at home, are uncertain about their future and suffer from bereavement following the loss of their biological parents [10–14]. ALHIV frequently experience verbal abuse, bullying, violence, stigma, and discrimination,[14–20] often internalize stigma and suffer from self-blame [13,21]. Lastly, ALHIV have to cope with issues related to the disclosure of their HIV status and the burden of lifelong HIV treatment [12,22,23].

There are little data on the prevalence of mental health problems among ALHIV in low- and middle-income countries. Studies from high-income countries estimate the prevalence of depression and anxiety disorders among ALHIV at about 25% [10]. For low- and middle-income countries data on the prevalence of mental disorders among ALHIV are sparse [10,24].

Mental health problems can adversely affect adherence to antiretroviral therapy (ART) and may lead to poor HIV treatment outcomes [25]. However, evidence on associations between mental health and markers of HIV treatment response among ALHIV is inconsistent. Some studies reported associations between mental health problems and low CD4 percentage, higher rates of unsuppressed HIV viral load, and HIV disease progression [10]. Still other studies found no evidence of such associations [10].

In low- and middle-income countries capacity to diagnose and manage mental disorders is limited [26]. The South African national treatment guidelines recommend routine mental health screening as part of adherence support [27], but the implementation of mental health services in ART programs is inconsistent [28]. Routine mental health screening is rarely done in public-sector ART programs in South Africa and the vast majority of ART patients affected by mental health problems remain undiagnosed and untreated [29].

We implemented routine mental health screening at a large pediatric HIV clinic in Johannesburg, South Africa to improve the identification and management of ALHIV affected by mental health problems. In this report, we describe the prevalence of and factors associated with positive screening for depression, suicidality, anxiety, trauma, and substance use.

## Methods

### Study design

We conducted a cohort study of HIV-positive adolescents who enrolled in HIV care at the Rahima Moosa Mother and Child Hospital in Johannesburg, South Africa. The clinic participates in the International epidemiology Databases to Evaluate AIDS (IeDEA) and regularly transfers routinely collected de-identified clinical data of HIV-positive mothers, children and adolescents to IeDEA data centres at the University of Cape Town and the University of Bern.

### Setting and participants

Rahima Moosa Mother and Child Hospital is an academic, public hospital. The clinic is the second-largest ART clinic for the treatment of children and adolescents living with HIV in Gauteng province. We included ALHIV, aged 10-19 years, who received ART at Rahima Moosa Mother and Child Hospital and participated at least once in mental health screenings during the study period from February 1, 2018, to January 1, 2020.

### Screening procedure

We offered adolescents aged 10-19 years old routine mental health screening at every three-monthly follow-up visit to the clinic. The screening consisted of a pre-screen for depression, anxiety, trauma, and substance use, followed by full-screen among ALHIV with a positive pre-screen (<u>Table 1</u>). The full-screen included the Patient Health Questionnaire (PHQ-9), the Generalized Anxiety Disorder 7 item (GAD-7) scale, the Primary Care PTSD Screen for DSM-5 (PC-PTSD-5) and the CAGE Adapted to Include Drugs (CAGE-AID) questionnaire (<u>Table 1</u>) [33–36]. Adolescents who screened positive for depression in the pre-screening also received a full screening for suicidality using the three-item screening tool from the Adolescent Innovations Project (AIP) working with Adolescents living with HIV handbook [37]. Those who reported past or current thoughts of self-harm or suicide or reported a previous suicide attempt screened positive in the full screening for suicidality. A PHQ-9 or GAD-7 score of 10 or higher was considered a positive full screen for depression and anxiety, respectively. For trauma, a PC-PTSD-5 score of 3 or higher and for substance use a CAGE-AID score of 2 or higher were considered positive full screens. In addition to mental health questions, screenings included six basic questions on food insecurity, assault, household conflicts, and socioeconomic situation. Counsellors (68.2 %, 3,044/ 4,461), and nurses (25.4%, 1,131) conducted most screens. Few screens (0.7%, 33) were conducted by doctors, social workers, or psychologists and the information on the screener was missing for 5.7% (253) of the screens. Adolescents who screened positive in a full-screen were referred and assessed by a senior doctor and social worker within the clinic and directed towards conclusive management which included referrals for counselling by on-site psychologists, peer-support, family meetings, and adherence counselling.

**Table 1.** Mental health and substance use screening model.

| Condition | Question | Pre-screen |  | Full-screen |  |
| --- | --- | --- | --- | --- | --- |
|  |  | Answers (points) | Positive screen | Tool | Positive screen |
| Depression | How often do you feel sad? | Every day (4)<br>Most of the time: 5-6 days a week (3)<br>Sometimes: 3-4 days a week (2)<br>Not much: 1-2 days a week (1)<br>Never (0) | 2 or higher | Screening for depression using the Patient Health Questionnaire for Adolescents (PHQ-9) [33] | 10 or higher |
| Suicidality |  |  | 2 or higher on depression pre-screen | Screening for suicidality using the Adolescent Innovations Project (AIP) working with Adolescents living with HIV handbook [37] | Past or current thoughts of self-harm/suicide or previous suicide attempt |
| Anxiety | How often do you feel worried? | Every day (4)<br>Most of the time: 5-6 days a week (3)<br>Sometimes: 3-4 days a week (2)<br>Not much: 1-2 days a week (1)<br>Never (0) | 2 or higher | Screening for anxiety using the Generalized Anxiety Disorder 7 item (GAD-7) scale [34] | 10 or higher |
| Trauma | Have you ever / since your last visit had a bad experience where you were scared that you or someone you love would be seriously hurt or killed? | Yes<br>No | Yes | Screening for trauma using the Primary Care PTSD Screen for DSM-5 (PC-PTSD-5) [35] | 3 or higher |
| Substance use | Do you drink alcohol or use drugs? | Yes<br>No | Yes | Screening for substance use using the CAGE Adapted to Include Drugs (CAGE-AID) scale [36] | 2 or higher |

### Definitions

The final screening outcome was positive if an ALHIV screened positive in any full-screen and as negative if he or she was screened at least once and screened negative. We defined the CD4 count at the start of the study as the value closest to the beginning February 1, 2018, within one year prior, and one year after that date. The window for CD4 at ART initiation was six months prior and one month after ART initiation. We defined the unsuppressed viral load as one viral load above 400 copies/mL.

### Statistical analysis

We used descriptive statistics to examine the characteristics of adolescents stratified by screening outcome. We calculated percentages of patients who screened positive, at the first screen, or ever, in pre- and full screens for depression, suicidality, anxiety, trauma, substance use, or any condition. Full screens were done conditionally on a positive pre-screen. For proportion, the denominator was the total population pre-screened. We calculated the mean and standard deviation of the PHQ-9, GAD-7, PC-PTSD-5, and CAGE-AID scores of adolescents who had a positive full screen for the particular condition. We estimated percentage of adolescents who had a positive full screen for two conditions for each possible pair of conditions and plotted the results in a heat map table.

We calculated unadjusted (OR) and adjusted odds ratios (aOR) with 95% confidence intervals (CI) for factors associated with positive full screens using logistic regression. We considered sociodemographic and clinical characteristics, and factors describing the life circumstances of patients in univariable analysis. Variables associated with a positive screen at a significance level of α <0.2 in univariable analysis were considered in multivariable analysis. Variables not significant at α<0.05 in multivariable analysis were eliminated from the final model [38].

We calculated ORs and aORs with 95% CIs for associations between positive full screens at first screen and unsuppressed viral load using logistic regression. Viral load testing was routinely performed annually. Hence, viral load testing was not necessarily performed on the date of mental health screening. We restricted the analysis of associations between positive full screens and unsuppressed viral load to adolescents who had a viral load test performed either on the day of screening or at the visit before the screening (i.e. within 100 days before screening). We did not consider viral load tests performed after screening because intervention following a positive mental health screen might influence viral load suppression, and therefore, might distort associations between screening outcomes and unsuppressed viral load. For patients who had more than one viral load test performed within the 100 days before screening, we selected the test closest to the screening date. In multivariable analysis, we adjusted for gender, age at screening (10-12 years, 13-15 years, or 16-19 years), current regimen (non-nucleoside reverse transcriptase inhibitor [NNRTI]-based, protease inhibitor [PI]-based, or other), age at ART initiation (<2 years, 2-4 years, 5-9 years, or ten years or older), CD4 cell count at ART initiation (<100 cells/µl, 100-199 cells/µl, 200-349 cells/µl, 350-499 cells/µl, ≥ 500 cells/µl or unknown), and regimen at ART initiation (NNRTI-based, PI-based, or other). Adjustment variables were selected a priori. In sensitivity analysis, we assessed associations of positive mental and substance use screens and unsuppressed viral load at a threshold of above 1000 copies/mL. Statistical analysis was done in Stata (Version 16, Stata Corporation, College Station, TX).

### Ethical considerations

The clinic has institutional ethical approval for the contribution of data to IeDEA. The Human Research Ethics Committee of the University of Cape Town, SA and the Cantonal Ethics Committee, Bern, Switzerland granted permission for analysis of this database. Institutional review boards have granted waivers of informed consent as the analyses use only de-identified data that are collected as part of routine patient care.

## Results

### Screening coverage

During the study period, 1,203 ART patients aged 10-19 visited the clinic of whom 1,088 (90.4%) were screened and included in our study. The reasons for omitting screening in 115 ALHIV (9.6%) were not documented. On average, patients participating in screenings were screened 8 times (IQR 6-9) over a median duration of 518 days (IQR 420-576) between first and last screening.

### Characteristics of adolescents screened for mental health and substance use problems

Half of the 1,088 ALHIV screened were male (50.5%). At the beginning of the study period, the median age of the study population was 13 years (IQR 10-15), patients were receiving ART for a median duration of 9 years (IQR 6-11 years). At ART initiation, the median age of patients was 3 years (IQR 1-7), median CD4 cell count was 496 cells/µl (IQR 262-853) (<u>Table 2</u>). Few adolescents (5.9%) reported that they had experienced food insecurity, 15.3% reported that they had experienced physical violence, and 14.1% reported a current or past conflict at home (<u>Table 3</u>).

**Table 2.** Demographic and clinical characteristics of adolescents screened for mental health and substance use problems in a paediatric HIV clinic in South Africa.

|  | Final screening outcome, n (%) |  |  |
| --- | --- | --- | --- |
|  | Negative | Positive | Total, n (%) |
|  | N=991 | N=97 | N=1,088 |
| **Characteristics at beginning of the study<sup>a</sup>** |  |  |  |
| Gender | 991 (100.0) | 97 (100.0) | 1,088 (100.0) |
| Male | 498 (50.3) | 51 (52.6) | 549 (50.5) |
| Female | 493 (49.7) | 46 (47.4) | 539 (49.5) |
| Years on ART | 991 (100.0) | 97 (100.0) | 1,088 (100.0) |
| 0-5 | 193 (19.5) | 12 (12.4) | 205 (18.8) |
| 6-10 | 527 (53.2) | 45 (46.4) | 572 (52.6) |
| >10 | 271 (27.3) | 40 (41.2) | 311 (28.6) |
| Median (IQR) | 9 (6-11) | 10 (7-12) | 9 (6-11) |
| Regimen | 991 (100.0) | 97 (100.0) | 1,088 (100.0) |
| NNRTI-based | 640 (64.6) | 59 (60.8) | 699 (64.2) |
| PI-based | 336 (33.9) | 38 (39.2) | 374 (34.4) |
| Other | 15 (1.5) | 0 (0.0) | 15 (1.4) |
| Age | 991 (100.0) | 97 (100.0) | 1,088 (100.0) |
| 9-12 | 522 (52.7) | 16 (16.5) | 538 (49.4) |
| 13-15 | 304 (30.7) | 25 (25.8) | 329 (30.2) |
| 16-19 | 165 (16.6) | 56 (57.7) | 221 (20.3) |
| Median (IQR) | 12 (10-14) | 16 (14-17) | 13 (10-15) |
| CD4 count in cells/μl | 991 (100.0) | 97 (100.0) | 1,088 (100.0) |
| <350 | 72 (7.3) | 13 (13.4) | 85 (7.8) |
| 350-499 | 104 (10.5) | 18 (18.6) | 122 (11.2) |
| 500-749 | 275 (27.7) | 27 (27.8) | 302 (27.8) |
| 750+ | 469 (47.3) | 33 (34.0) | 502 (46.1) |
| Unknown | 71 (7.2) | 6 (6.2) | 77 (7.1) |
| Median (IQR) | 766 (545-990) | 671 (397-840) | 747 (537-982) |
| **Characteristics at ART initiation** |  |  |  |
| Regimen, n (%) | 991 (100.0) | 97 (100.0) | 1,088 (100.0) |
| NNRTI-based | 562 (56.7) | 73 (75.3) | 635 (58.4) |
| PI-based | 379 (38.2) | 23 (23.7) | 402 (36.9) |
| Other | 50 (5.0) | 1 (1.0) | 51 (4.7) |
| Age, n (%) | 991 (100.0) | 97 (100.0) | 1,088 (100.0) |
| <2 | 386 (39.0) | 15 (15.5) | 401 (36.9) |
| 2-4 | 225 (22.7) | 29 (29.9) | 254 (23.3) |
| 5-9 | 279 (28.2) | 40 (41.2) | 319 (29.3) |
| 10-19 | 101 (10.2) | 13 (13.4) | 114 (10.5) |
| Median (IQR) | 3 (1-6) | 5 (3-8) | 3 (1-7) |
| CD4 count in cells/μl, n (%) | 991 (100.0) | 97 (100.0) | 1,088 (100.0) |
| <100 | 72 (7.3) | 9 (9.3) | 81 (7.4) |
| 100-199 | 38 (3.8) | 7 (7.2) | 45 (4.1) |
| 200-349 | 92 (9.3) | 11 (11.3) | 103 (9.5) |
| 350-499 | 92 (9.3) | 9 (9.3) | 101 (9.3) |
| 500+ | 302 (30.5) | 24 (24.7) | 326 (30.0) |
| Unknown | 395 (39.9) | 37 (38.1) | 432 (39.7) |
| Median (IQR) | 505 (271-873) | 410 (186-648) | 496 (262-853) |

Data are n (%) unless otherwise specified.
<sup>a</sup> Beginning of study period was February 01, 2018.
Abbreviations: ART, antiretroviral therapy; NNRTI, non-nucleoside reverse transcriptase inhibitors; PI, protease inhibitors; IQR, interquartile range

**Table 3.** Life circumstances of adolescents screened for mental health and substance use problems at a paediatric HIV clinic in South Africa.

|  | Final screening outcome, n (%) |  |  |
| --- | --- | --- | --- |
|  | Negative | Positive | Total, n (%) |
|  | N=991 | N=97 | N=1,088 |
| Experienced food insecurity | 991 (100.0) | 97 (100.0) | 1,088 (100.0) |
| Never | 936 (94.5) | 87 (89.7) | 1,023 (94.0) |
| Currently or in the past | 54 (5.4) | 10 (10.3) | 64 (5.9) |
| Unknown/refused/missing | 1 (0.1) | 0 (0.0) | 1 (0.1) |
| Experienced physical violence | 991 (100.0) | 97 (100.0) | 1,088 (100.0) |
| Never | 844 (85.2) | 75 (77.3) | 919 (84.5) |
| Currently or in the past | 146 (14.7) | 21 (21.6) | 167 (15.3) |
| Unknown/refused/missing | 1 (0.1) | 1 (1.0) | 2 (0.2) |
| Conflict in household | 991 (100.0) | 97 (100.0) | 1,088 (100.0) |
| Never | 863 (87.1) | 71 (73.2) | 934 (85.8) |
| Currently or in the past | 127 (12.8) | 26 (26.8) | 153 (14.1) |
| Unknown/refused/missing | 1 (0.1) | 0 (0.0) | 1 (0.1) |
| Employment of household member | 991 (100.0) | 97 (100.0) | 1,088 (100.0) |
| Never | 35 (3.5) | 2 (2.1) | 37 (3.4) |
| Currently or in the past | 955 (96.4) | 95 (97.9) | 1,050 (96.5) |
| Unknown/refused/missing | 1 (0.1) | 0 (0.0) | 1 (0.1) |
| Household grant | 991 (100.0) | 97 (100.0) | 1,088 (100.0) |
| Never | 190 (19.2) | 24 (24.7) | 214 (19.7) |
| Currently or in the past | 745 (75.2) | 72 (74.2) | 817 (75.1) |
| Unknown/refused/missing | 56 (5.7) | 1 (1.0) | 57 (5.2) |

Data are n (%).

### Screening outcomes

Out of all adolescents who were pre-screened, 6.9% had a positive full screen for any condition at the first screen: 3.6% for depression, 2.2% for suicidal concern, 1.9% for anxiety, 2.2% for trauma, and 0.9% for alcohol/substance abuse. The prevalence of positive full-screens increased with age (<u>Table 4</u>). In repeated screening, few additional adolescents screened positive in full-screens. In total, 8.9% of pre-screened adolescents ever screened positive in a full-screen for any condition: 4.4% for depression, 2.7% for suicidal concern, 2.2% for anxiety, 3.5% for trauma, and 1.7% for alcohol/substance abuse (<u>Table 4</u>).

**Table 4.**
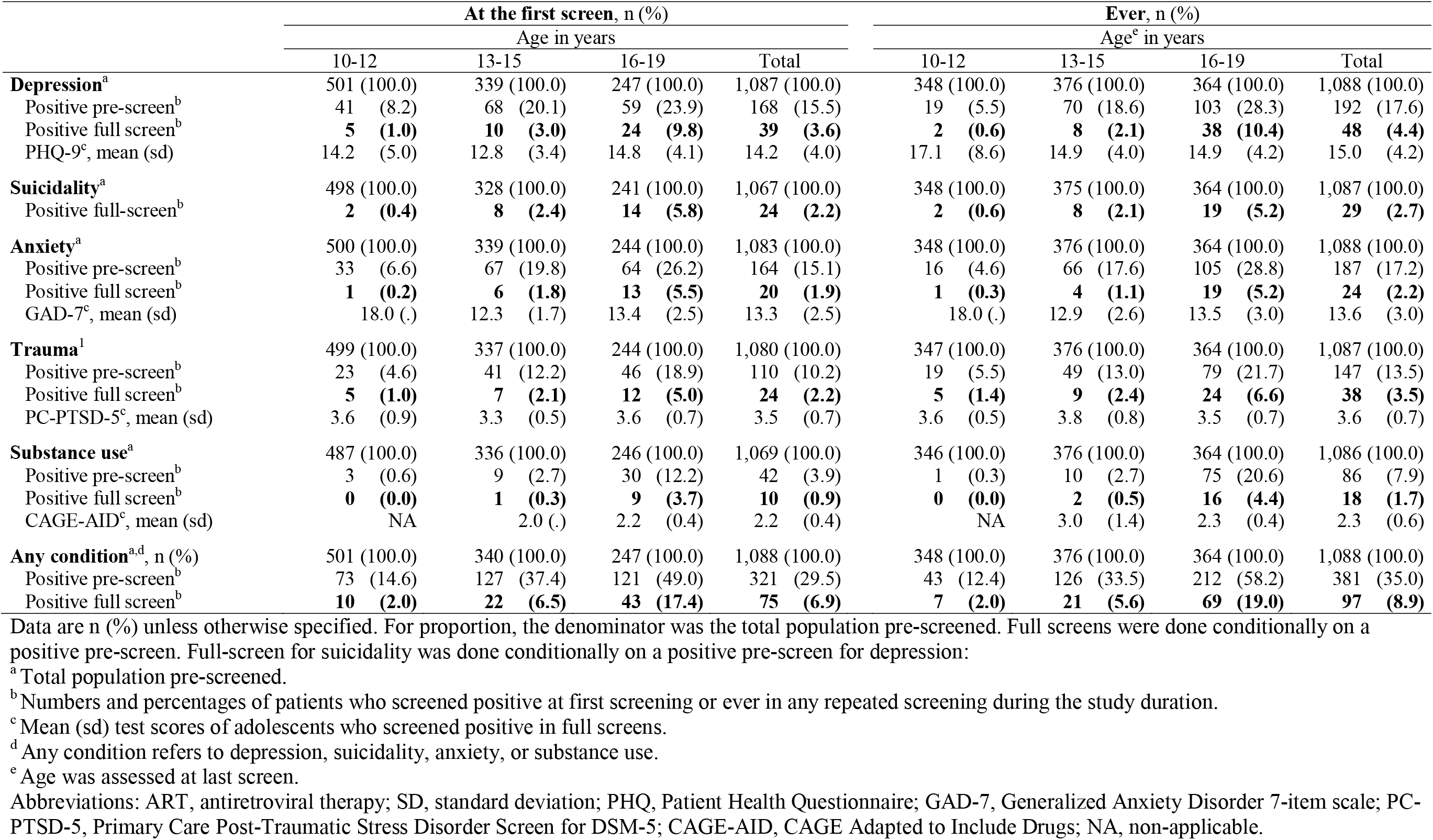
Mental health and substance use screening outcomes of adolescents attending a paediatric HIV clinic in South Africa.

### Concurrence of multiple mental health and substance use problems

Positive full-screens for multiple conditions were common: among the ALHIV who had a positive full-screen for one condition, 36.1% (35/97) had another positive full-screen and 19.6% (19/97) had positive full-screens for two additional conditions. <u>Figure 1</u> shows the concurrence of positive full-screens for each possible pair of conditions. Positive full-screens for depression and suicidality often concurred: 39.6% (19/48) of adolescents who screened positive for depression also screened positive for suicidality. Other common concurrences include positive depression screens among adolescents with anxiety (45.8%, 11/24) or trauma (39.5%, 15/38).

**Figure 1.**
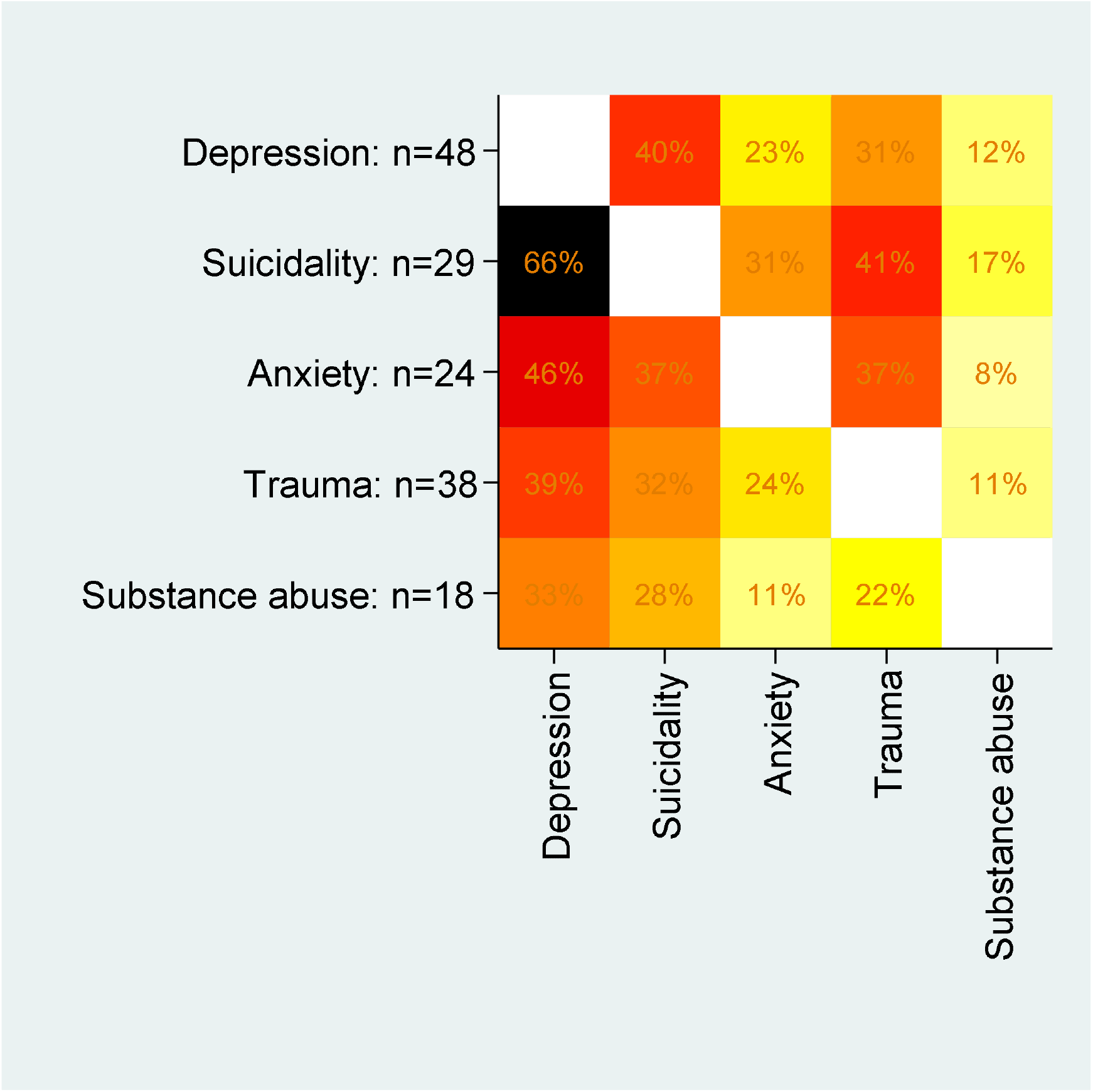
Concurrence of positive full screen for mental health and substance use conditions. Percentage of adolescents who had a positive full screen for the condition shown on the x-axis among those who had a positive full screen for the condition shown on the y-axis. N represents the denominator. Darker colours represent higher values.

### Patient characteristics associated with positive full screens

Older age was associated with higher odds for positive screening for all five conditions: aORs for positive screening for any condition for adolescents 16-19 years old compared to those 10-12 years were 11.46 (95 CI 5.17-25.40) (<u>Table 5</u>). There were no gender differences in the odds of positive screening for depression, suicidality, anxiety, or trauma. However, female gender was strongly associated with decreased odds for a positive substance use screen (aOR 0.19, 95% CI 0.05-0.66). Experience of physical violence was associated with positive full screen for suicidality, anxiety, trauma, and any condition and household conflicts were associated with positive full screen for depression, and any condition (<u>Table 5</u>).

**Table 5.**
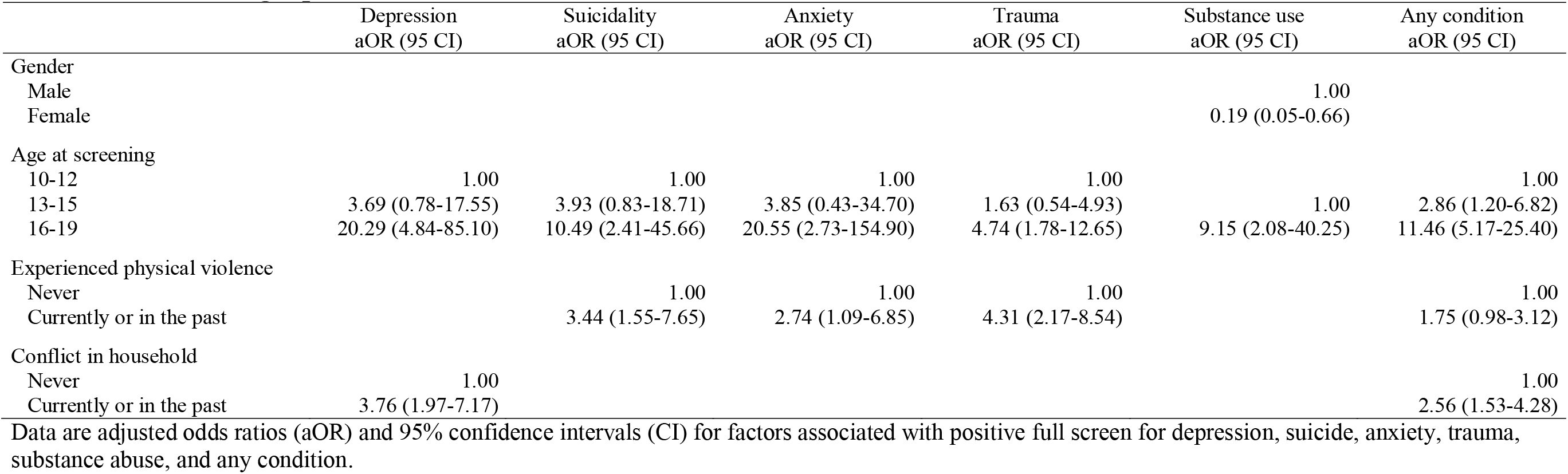
Patient characteristics associated with positive full screens for mental health and substance use problems among adolescents attending a paediatric HIV in South Africa.

|  | Depression<br>aOR (95 CI) | Suicidality<br>aOR (95 CI) | Anxiety<br>aOR (95 CI) | Trauma<br>aOR (95 CI) | Substance use<br>aOR (95 CI) | Any condition<br>aOR (95 CI) |
| --- | --- | --- | --- | --- | --- | --- |
| Gender |  |  |  |  |  |  |
| Male |  |  |  |  | 1.00 |  |
| Female |  |  |  |  | 0.19 (0.05-0.66) |  |
| Age at screening |  |  |  |  |  |  |
| 10-12 | 1.00 | 1.00 | 1.00 | 1.00 |  | 1.00 |
| 13-15 | 3.69 (0.78-17.55) | 3.93 (0.83-18.71) | 3.85 (0.43-34.70) | 1.63 (0.54-4.93) | 1.00 | 2.86 (1.20-6.82) |
| 16-19 | 20.29 (4.84-85.10) | 10.49 (2.41-45.66) | 20.55 (2.73-154.90) | 4.74 (1.78-12.65) | 9.15 (2.08-40.25) | 11.46 (5.17-25.40) |
| Experienced physical violence |  |  |  |  |  |  |
| Never |  | 1.00 | 1.00 | 1.00 |  | 1.00 |
| Currently or in the past |  | 3.44 (1.55-7.65) | 2.74 (1.09-6.85) | 4.31 (2.17-8.54) |  | 1.75 (0.98-3.12) |
| Conflict in household |  |  |  |  |  |  |
| Never | 1.00 |  |  |  |  | 1.00 |
| Currently or in the past | 3.76 (1.97-7.17) |  |  |  |  | 2.56 (1.53-4.28) |

Data are adjusted odds ratios (aOR) and 95% confidence intervals (CI) for factors associated with positive full screen for depression, suicide, anxiety, trauma, substance abuse, and any condition.

### Associations between positive full screens and unsuppressed viral load

Three-quarters of the adolescents screened for mental health, and substance use problems (74.6%, 812/1,088) had a viral load test performed on the day of or within 100 days before mental health screening. They were included in the analysis of associations between positive mental health screens and unsuppressed viral load. The median time between viral load test and mental health screening among adolescents included in this analysis was 0 days (IQR 0-84 days). The prevalence of unsuppressed viral load was 17.4% (131/753) in adolescents who screened negative for all conditions, and 33.9% (20/59) in adolescents who screened positive for at least one condition. In unadjusted analysis, positive full-screens for depression (OR 2.56, 95% CI 1.16-5.67), suicidality (OR 3.02, 95% CI 1.06-8.62), trauma (OR 3.51, 95% CI 1.29-9.59), substance use (OR 9.03, 95% CI 2.23-36.53), or any condition (OR 2.43, 95% CI 1.38-4.31) were associated with unsuppressed viral load (<u>Figure 2</u>). In adjusted analysis, positive full screen for depression (aOR 2.39, 95% CI 1.02-5.62), trauma (aOR 3.18, 95% CI 1.11-9.07), substance use (aOR 7.13, 95% CI 1.60-31.86), or any condition (aOR 2.17, 95% CI 1.17-4.02) remained associated with higher odds of unsuppressed viral load (<u>Figure 2</u>). Results were similar in the sensitivity analysis in which we used a threshold of 1000 copies per millilitre to define unsuppressed viral load (<u>Figure S1</u>). Positive screening for any condition remained associated with higher odds of unsuppressed viral load at the threshold of above 1000 copies per millilitre in unadjusted (OR 2.77, 95% CI 1.53-5.01) and adjusted sensitivity analysis (aOR 2.36, 95% CI 1.25-4.47) (<u>Figure S1</u>).

**Figure 2.**
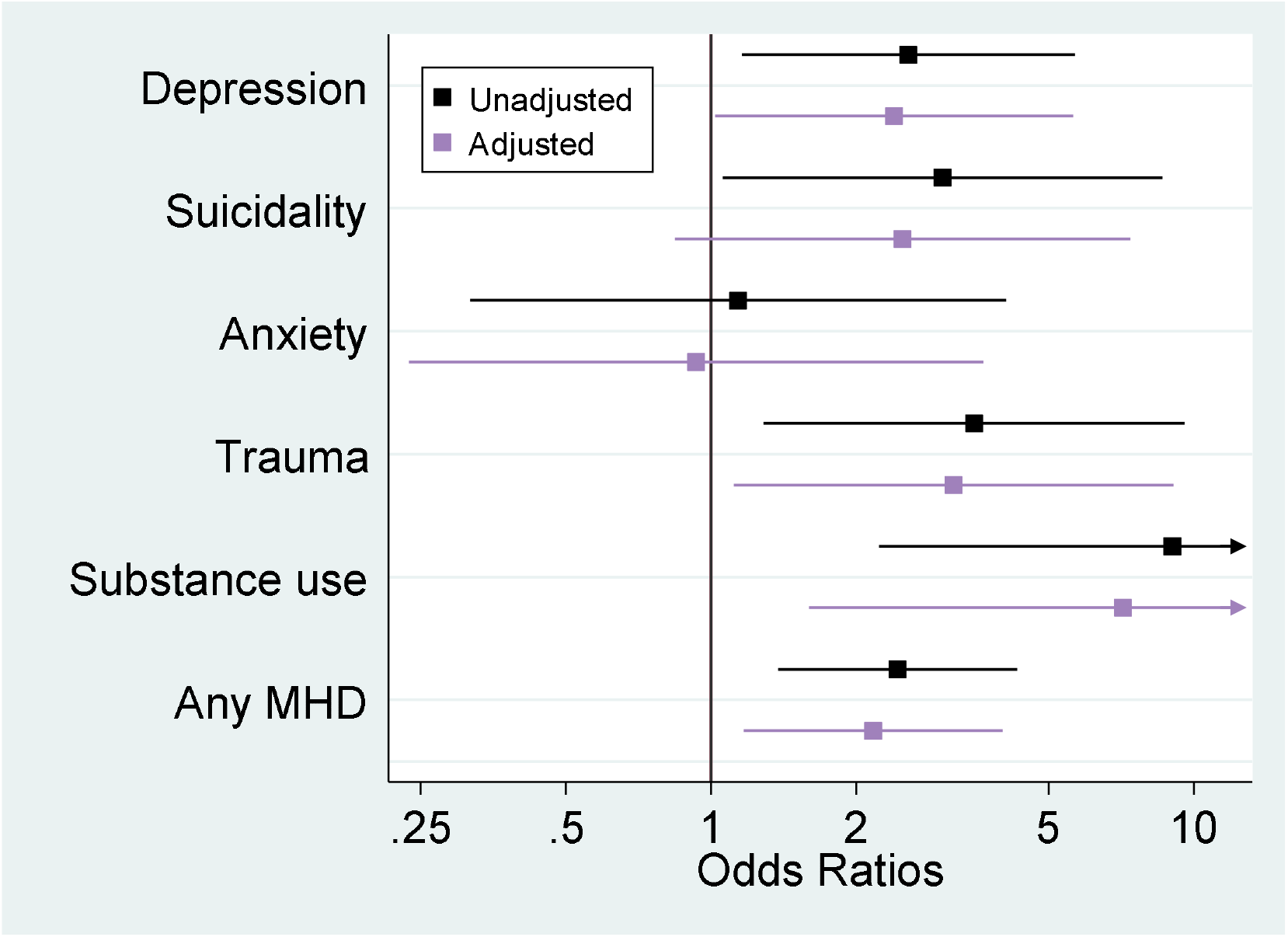
Associations between positive full screens for mental health and substance use problems and unsuppressed viral load. Adjusted and unadjusted odds ratios and 95% confidence intervals for associations between positive full-screens for depression, suicidality, anxiety, trauma, substance use, and any condition and unsuppressed viral load (i.e. viral load above 400 copies/mL. Odds ratios were adjusted for gender, age at screening, current regimen, age, CD4 cell count, and regimen at ART initiation.

## Discussion

Overall, about 9% of ALHIV screened positive for mental health or substance use problems with a much higher prevalence among adolescents aged 16-19 years (19%) than among those aged 10-12 years (2%). Depression was the most common mental health problem. Only a few adolescents screened positive for substance abuse. Many adolescents screened positive for more than one conditions. Older age, the experience of physical violence, and household conflicts were associated with positive mental health screens. One-third of adolescents who screened positive for a mental health or substance use problem had an unsuppressed viral load. A positive screen for mental health or substance use problems was strongly associated with unsuppressed viral load.

Our study has several strengths. We conducted our study in a large public ART clinic in South Africa. The majority of HIV-positive adolescents live in sub-Saharan Africa and receive care in a similar health care setting. Participants enrolled in our study received only routine services that are the standard of care for adolescent ART patients in South Africa. The study setting and our “naturalistic” approach contribute to the external validity of our study and make our results applicable to a large proportion of the ALHIV globally. Our analyses were adequately powered to examine risk factors associated with mental health and substance use problems. The longitudinal study design allowed for repeated mental health screening and we could assess the prevalence of positive mental health screens over a period of two years.

Our results have to be interpreted in light of several limitations. Although we used validated screening tools [33–36,39,40], we did not validate our two-stage screening model. The prevalence of positive screening tests in our study was relatively low, and it is possible that we missed adolescents with mental health or substance use problems during the pre-screening phase. We used data from routinely performed viral load tests to examine associations between mental health and unsuppressed viral load. One-quarter of patients could not be included in this analysis because they had no recent viral load test result available. For one-quarter of patients, we used a viral load result from a test that was performed about 3 months before mental health screening. Changes in viral load occurring between viral load testing and mental health screening could distort associations between positive screens and unsuppressed viral load.

The prevalence of mental health and substance use problems observed in this study was lower than in other studies. It is estimated that mental health problems affect about 10-20% of children and adolescents worldwide [41,42]. In ALHIV, the prevalence of mental health problems is substantially higher [10]. In the United States, more than half of the ALHIV who enrolled in two major HIV cohort studies had a diagnosis of a psychiatric disorder [43–45]. In another large cohort study which recruited participants from 29 US sites, 61% of the ALHIV showed psychiatric symptoms [46]. A cross-sectional survey conducted in Soweto Township in Johannesburg, South Africa found that one-third of the adolescents recruited at an average age of 16 years had symptoms of moderate to severe depression. One-fifth of the adolescents reported substance use problems [47]. Another study conducted at five paediatric ART clinics in Johannesburg reported that 27% of ALHIV showed symptoms for depression, anxiety, or post-traumatic stress disorder and 24% reported suicidality [14]. Similar estimates were reported in studies from other Southern African countries. In Malawi, 19% of the adolescents attending two large urban ART clinics (mean age 14.5 years) were diagnosed with depression [48]. In Zambia, 25% of adolescents aged 15-19 enrolled in HIV care at a large in Lusaka screened positive for depression. In Uganda, 46% of adolescents (10-19 years) had depressive symptoms [49,50].

Although estimates of the prevalence of mental health problems among ALHIV are not directly comparable, given differences in study setting, age range of study populations, and instruments used to measure mental health problems, the proportion of adolescents positive mental health screen in our study appears to be below the expected prevalence in this population. Low sensitivity of the two-stage screening approach used in our study is one possible explanation for the lower than expected prevalence.

Our study provides strong evidence for associations between mental health and substance use problems and unsuppressed viral load. Existing evidence on this association is contradictory [10].

In a longitudinal study of children and adolescents living with HIV from the US and Puerto Rico, those with symptoms of anxiety had a 40% lower odds of having unsuppressed viral load (viral load >400 copies/mL). Symptoms of attention deficit hyperactivity disorder, disruptive behaviour or depression were not associated with unsuppressed viral load [51]. A cohort study of children and ALHIV from Botswana those who scored high for cognitive, emotional, and behavioural problems were more likely to have an unsuppressed viral load (viral load >400 copies/mL) than those with a lower score [52]. In Windhoek, Namibia, a study found no evidence for associations between mental health problems and viral load suppression [53]. In a study from New York City, disruptive behaviour disorder was strongly associated with unsuppressed viral load at a threshold of greater than >1000 copies per millilitre. By contrast, two other studies from New York found no evidence for associations between mental disorders and unsuppressed viral load [44,54].

Our research has several implications for policy and practice. We showed that ALHIV who are affected by mental health or substance use problems are a highly vulnerable population, who merit specific clinical attention. Our study demonstrates that the integration of routine screening and management of mental health and substance use problems in a high-burden public-sector clinic in South Africa is feasible. Our experience shows that the response to mental health conditions requires a tailored response, adapted to the setting and population of ALHIV. While few adolescents with severe mental health problems need a referral to specialized mental health services, referrals to services outside of the clinic are often not practical, and many mental health problems can be addressed by the ART clinic. We piloted a range of interventions including family meetings, referral to on-site psychologists, social workers, and peer-support groups. Common underlying causes of mental health problems, including bullying, conflicts in the home, or food insecurity could be identified and addressed with these interventions. While screening and management of mental and substance use problems has the potential to improve adherence, virologic response, and quality of life of ALHIV, we could not evaluate the effectiveness of the piloted interventions.

## Conclusions

HIV-positive adolescents who are affected by mental health or substance use problems are a highly vulnerable population, who merit specific clinical attention. More work is needed to develop and evaluate strategies for screening and management of mental health and substance use problems in a paediatric ART programs in low- and middle-income countries.

## Data Availability

All data were obtained from the International epidemiology Database to Evaluate AIDS Southern Africa collaboration (IeDEA-SA, https://www.iedea-sa.org/). Data cannot be made available online because of legal and ethical restrictions. To request data, readers may contact IeDEA for consideration by filling out the online form available at https://www.iedea-sa.org/contact-us/.

## Authors’ contributions

AH, KT, MD, and ME conceptualized and designed the study. SP developed the screening model. KT, SP, KB, MM, GS, SS, PvG, and MT assisted in the implementation, fieldwork, or data collection. AH conducted the statistical analysis and wrote the first draft of the manuscript. NM assisted with data extraction and curation of the database. All authors contributed to interpretation of data and provided critical inputs in the draft manuscript. All authors have read and approved the final manuscript.

## Acknowledgements

We thank the adolescents whose data were used in this analysis, as well as their caregivers. We also thank the IeDEALJSA Data Centre teams at the Universities of Cape Town and Bern.

---

**Figure S1.**
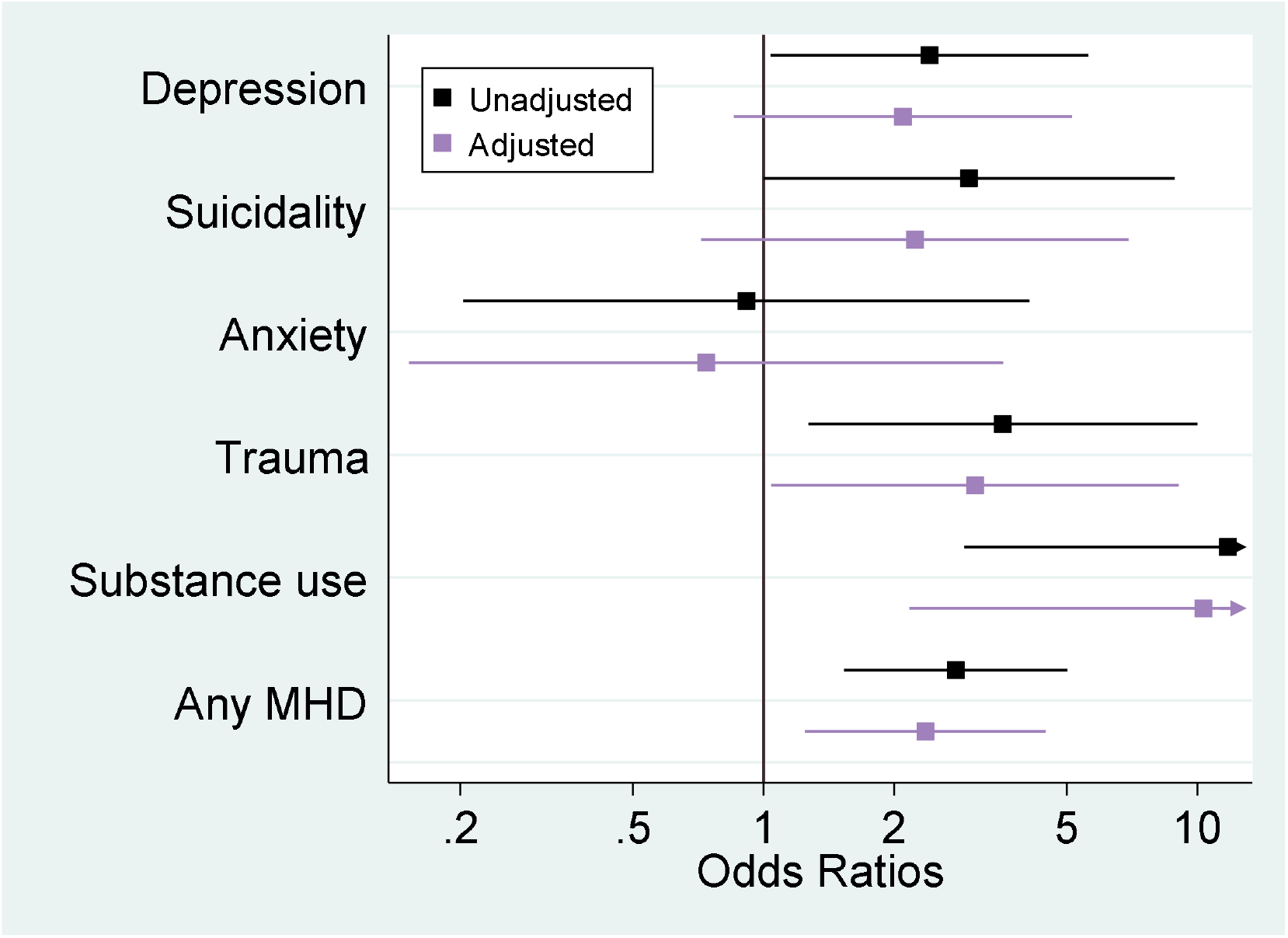
Sensitivity analysis of associations between positive full screens for mental health or substance use problems and viral load >1000 copies/mL. Adjusted and unadjusted odds ratios and 95% confidence intervals for associations between positive full-screens for depression, suicidality, anxiety, trauma, substance use, and any condition and viral load above 1000 copies/mL. Odds ratios were adjusted for gender, age at screening, current regimen, age, CD4 cell count, and regimen at ART initiation.

